# Convalescent plasma treatment of B-cell depleted patients with COVID-19: systematic review and individual participant data meta-analysis

**DOI:** 10.1101/2025.05.15.25327576

**Authors:** Solomiia Zaremba, Daniele Focosi, Wyatt W. Pruter, Massimo Franchini, Diana B. Collantes Hoyos, Mario Cruciani, Alex J. Miller, Juan G. Ripoll, Arturo Casadevall, Sidna M. Tulledge-Scheitel, Nathalie Rufer, Thomas Hueso, Justin E. Juskewitch, Camille M. van Buskirk, Petros Ioannou, Francesco Lanza, Raymund R. Razonable, Ferenc Magyari, László Imre Pinczés, Ravindra Ganesh, Claudia M. Denkinger, Ryan T. Hurt, Maike Janssen, James R. Stubbs, Carsten Müller-Tidow, Jeffrey L. Winters, Karin Holm, Sameer A. Parikh, Gordana Simeunovic, Neil E. Kay, Bart Rijnders, Scott R. Wright, Nahema Issa, Hél□ne Chaussade, Rickey E. Carter, Darrell R. Schroeder, Jonathon W. Senefeld, Michael J. Joyner

## Abstract

**Background:** COVID-19 convalescent plasma (CCP) is obtained from people recently recovered from COVID-19 and contains viral-neutralizing antibodies. Because such treatment is safe and effective against SARS-CoV-2, the US Food and Drug Administration (FDA) has recently authorized the use of CCP for COVID-19 patients with immunodeficiencies. Currently available CCP is a “hybrid” product with antibodies from individuals who had both infection and vaccination (vaccine-boosted CCP). In this context, there is growing interest in CCP treatment outcomes in specific groups of immunocompromised patients. Specifically, B-cell depleted patients are at risk of not producing antibodies after either infection or vaccination. Hence, B-cell depleted patients are conceivably among those who would benefit the most from CCP. We thus conducted a systematic review and individual patient data meta-analysis to assess characteristics associated with 60-day survival in B-cell depleted patients transfused with CCP.

**Methods:** The last search was April 2^nd^, 2024, and included all studies using CCP in B-cell depleted patients. Whenever not available in the publication, we requested individual participant data from corresponding authors of eligible studies. Risk of bias was assessed using Joanna Briggs Institute Critical Appraisal Tools. Data were analyzed using conditional logistic regression.

**Results:** We describe individual patient data extracted from 85 studies and synthesized into a cohort of 570 patients. The overall 60-day survival rate was 86.5%. Of patients with information available, 70.1% achieved SARS-CoV-2 clearance, and 64.3% had clinical improvement within 5 days of CCP transfusion. After controlling for age, sex, calendar year of infection and World Health Organization (WHO) disease severity, we found a significant association between 60-day survival and days since last anti-CD20 dose (OR=1.16 per 10-day increase; 95% CI 1.04 to 1.29; p=0.007) and transfusion of vaccine-boosted CCP (OR=9.49; 95% CI 2.01 to 44.82; p=0.005), but not with concomitant remdesivir treatment (OR=1.31; 95% CI 0.66 to 2.61; p=0.440).

**Discussion:** Our study is limited to individual participant data analysis, with a majority of the studies included being case series and case reports. Overall survival in our cohort of B-cell depleted patients was consistent with prior meta-analysis of randomized controlled trials on survival of immunocompromised patient transfused with CCP (∼84%). A novel finding from this analysis is that vaccine-boosted CCP with a presumably higher content of neutralizing antibodies is associated with a high survival benefit.

**Registration:** The protocol for this systematic review and individual participant data meta-analysis was registered with PROSPERO (CRD42024516513) on March 1st, 2024.

## Introduction

Anti-CD20 monoclonal antibodies (e.g. rituximab) are commonly used for the treatment of patients with hematologic malignancies and autoimmune disorders.^1,2^ By selectively targeting B-lymphocytes, such treatments impair the host ability to produce infection-neutralizing antibodies. Full recovery of the B-cell pool to normal levels can take up to 12 months after the last dose of anti-CD20 therapy.^3^ In this way, B-cell depleted patients are unlikely to mount robust endogenous antibody responses to either vaccine^4,5^ or natural infections, and thus are at higher risk of infections (including SARS-CoV-2 infection) and mortality. Indeed, treatment with anti-CD20 monoclonal antibodies has been shown to be associated with prolonged SARS-CoV-2 shedding.^6–10^ Persistent SARS-CoV-2 viremia in immunocompromised patients has also been shown to be associated with clinical severity of COVID-19 and mortality, highlighting the importance of prompt viral neutralization.^11^ Additionally, uncontrolled infection may lead to viral evolution and generation of novel SARS-CoV-2 variants in such patients.^12^

COVID-19 convalescent plasma (CCP) has a robust safety and efficacy profile against SARS-CoV-2 among patients with immunodeficiencies.^13–15^ CCP contains functional, polyclonal antibodies that can neutralize a broad range of SARS-CoV-2 variants and mutations, and leads to other non-neutralizing antibody-mediated effects and immunomodulation.^16,17^ Expert guidance on the use of CCP for immunocompromised patients includes anti-CD20 B-cell depleted patients (among others) as a target population recommended for CCP therapy^14^, because in such patients CCP transfusion can benefit both acute symptomatic SARS-CoV-2 infection and protracted COVID-19.

However, there remains a limited number of studies on the clinical efficacy of CCP in B-cell depleted patients. Because CCP provides the viral-neutralizing antibodies B-cell depleted patients are lacking, this patient population may be among those who benefit the most from such therapy. Thus, we aimed to describe characteristics and outcomes of B-cell depleted patients infected with SARS-CoV-2 and transfused with CCP, whose cases were reported in the literature. Our overall goal was to gain insight into the characteristics of the patients, CCP treatment timing, and CCP quality that might influence outcomes in immunocompromised patients.

Consequently, we conducted a systematic review and individual participant data (IPD) meta-analysis to summarize available reports on the use of CCP in patients with secondary immunodeficiency due to anti-CD20 B-cell depletion.

## Methods

### Protocol and registration

The protocol for this systematic review and individual participant data meta-analysis was registered with PROSPERO (CRD42024516513) on March 1st, 2024.

### Information sources and search

This systematic review and IPD meta-analysis aimed to evaluate outcomes of patients who have undergone B-cell depleting therapy with anti-CD20 medications and were transfused with CCP. Our review was conducted according to the guidance provided in the PRISMA-IPD (Preferred Reporting Items for Systematic reviews and Meta-Analyses, Table S10). A previously published systematic review on the impact of CCP transfusion on COVID-19 mortality in patients with immunodeficiencies^15^ was used to identify potentially eligible studies. That review included studies published between January 1^st^ 2020 and August 12^th^ 2022. Sources for the search included PubMed, MEDLINE, Google Scholar and medRχiv. An additional search to update the results was conducted between August 12^th^ 2022 and April 2^nd^ 2024. Keywords and Medical Subject Heading (MeSH) terms used for the search were similar to those used in the previous systematic review, although more specific to the type of immunodeficiency: (*COVID-19* OR *SARS-CoV-2* OR *coronavirus disease 2019*) AND (*convalescent plasma* OR *immune plasma* OR *hyperimmune plasma*) AND (*immunosuppression* OR *immunodeficiency* OR *immunocompromised* OR *cancer* OR *malignancy* OR *hematological* OR *oncologic* OR *lymphoma* OR *leukemia* OR *myeloma* OR *multiple sclerosis* OR *agammaglobulinemia* OR *hypoglobulinemia* OR *autoimmune disorder* OR *BCDT* OR *B-cell depletion*). Full-text translation to English was required for the inclusion of the article.

This search yielded 850 studies, including 145 studies from the previous review^15^ and 705 studies from the additional search (Figure 1).

**Figure 1.**
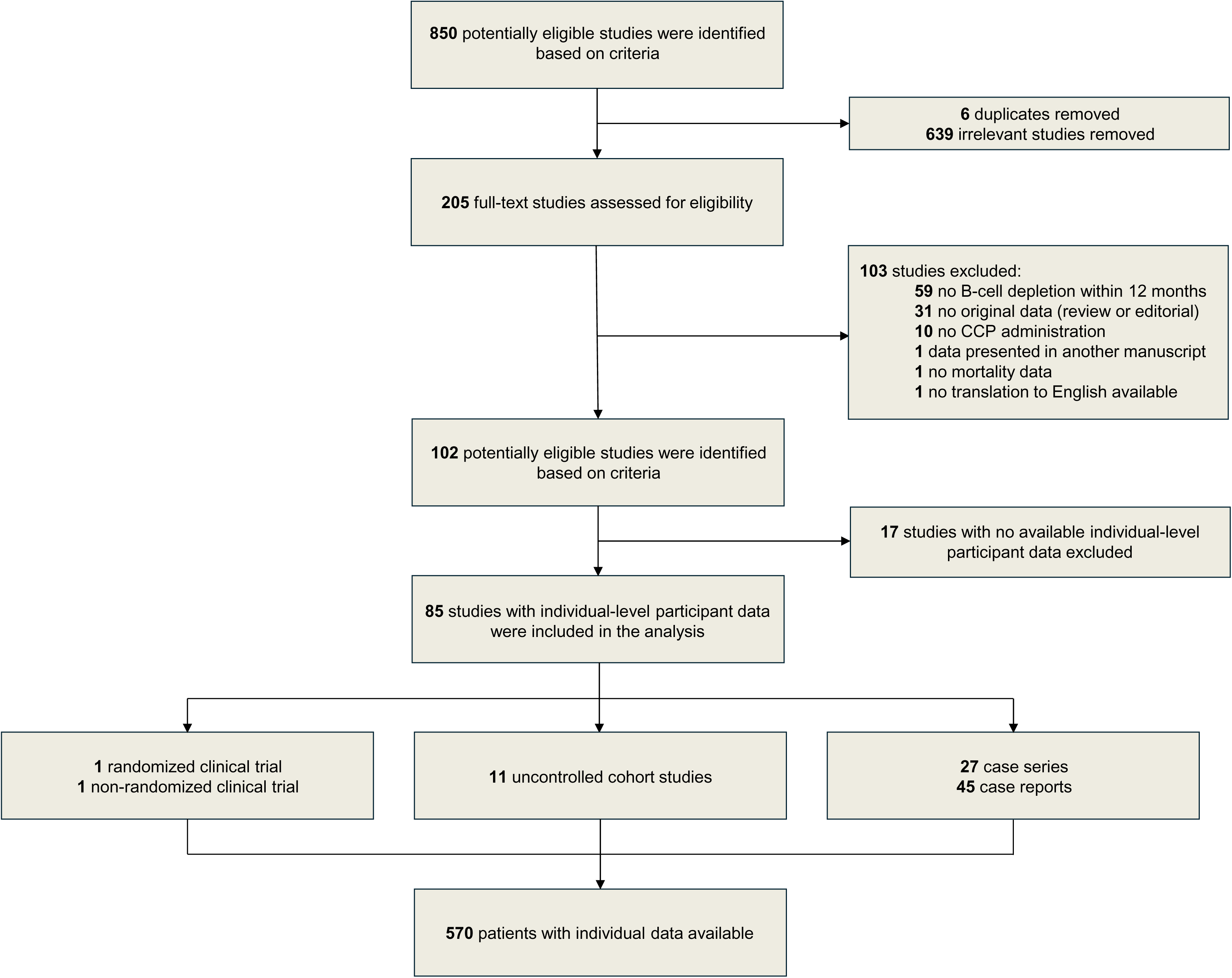
Flowchart of studies inclusion and exclusion criteria.

### Study selection and data collection processes

After initial screening of 850 potentially eligible manuscripts, we removed 639 irrelevant studies and six duplicates. 205 full-text manuscripts were then further assessed for eligibility. Studies that included only patients with primary immunodeficiencies were excluded. We then contacted the authors who reported patients with secondary immunodeficiencies transfused with CCP but did not specify the underlying cause of immunodeficiency or did not report individual patient-level data. This review did not have restrictions on study type, thus randomized controlled trials (RCT), non-RCT, cohort studies, case series and case reports were all eligible for inclusion. However, manuscripts with no original data (reviews and editorials) were excluded. Studies were excluded if IPD was unavailable, patients did not receive a B-cell depleting therapy within 12 months prior to COVID-19 diagnosis, if patients did not receive CCP treatment for COVID-19, if the study did not report our primary outcome of interest (i.e. mortality), if data were already included in another manuscript as part of larger cohort, and if no translation to English was available. The full-text screening process resulted in exclusion of 103 studies (Figure 1). Out of 102 potentially eligible studies, we received or abstracted individual patient data from 85 studies. Corresponding authors directly input IPD into a combined dataset for all eligible patients. For the studies that included IPD in primary text, two independent reviewers out of potentially available four reviewers extracted the data from each manuscript (D.F., S.Z., D.C.H., A.J.M.), then disagreements were resolved by third reviewer (J.W.S.).

Abstracted data included: patient age and sex, clinical status before CCP transfusion (8-point World Health Organization [WHO] COVID-19 disease severity score,^19^ need for mechanical ventilation, and serum anti-Spike antibody status), underlying condition and concomitant therapies, type(s) of B-cell depleting agents, CCP therapy (number of CCP units administered, volume of plasma per CCP unit, total volume of CCP transfused), use of vaccine-boosted CCP, time since the symptoms onset to first CCP transfusion, time since the first hospital admission to first CCP transfusion (if applicable), concomitant COVID-19 therapies (intravenous immunoglobulin, anti-spike monoclonal antibodies, steroids, remdesivir, nirmatrelvir/ritonavir, hydroxychloroquine), patient clearance of SARS-CoV-2, rapid clinical improvement (defined as symptoms improvement and/or reduction in supplemental oxygen requirement within 5 days of CCP transfusion), duration of follow-up in days, and all-cause mortality. We collected all data possible within the constraints of availability, as not all the data were available for each patient. Our primary outcome of interest was 60-day survival following CCP transfusion or initial CCP transfusion in the cases of multiple CCP transfusions. For this primary outcome of 60-day survival, patients who died but did not have a time to death indicated were assumed to have died prior to 60 days; and patients who were indicated to have survived but had follow-up of less than 60 days were assumed to have survived to 60 days.

### Eligibility criteria

Eligible participants were patients with secondary immunodeficiencies due to treatment with anti-CD20 monoclonal antibodies who were diagnosed with COVID-19 and were then transfused with CCP. Both hospitalized and non-hospitalized patients with COVID-19 were considered. Patients were excluded if the most recent B-cell depleting therapy was more than 12 months before COVID-19 diagnosis. No control participants were included in this systematic review and IPD meta-analysis. Participants were eligible for inclusion if the information on the primary outcome (60-day survival) was available. Participants who did not meet these criteria were excluded on the individual level. If no IPD were presented in the manuscript, studies with potentially eligible cohorts (e.g. COVID-19 patients with hematologic malignancies transfused with CCP, COVID-19 patients with autoimmune disorders transfused with CCP) were considered for inclusion. After obtaining further details on the patient’s underlying condition and treatment (anti-CD20 B-cell depletion status in question) from the corresponding authors, patients were included on the individual basis if fulfilled these criteria. Additionally, a study was considered eligible if at least one participant from the cohort met inclusion criteria. Studies were excluded if no information on anti-CD20 treatment was available.

### Ethical Approval and Quality Assessment

This study was exempt from institutional review board approval at Mayo Clinic, as only previously published data were included. Thus, patient informed consent was waived.

Joanna Briggs Institute (JBI) Critical Appraisal Tools were used for risk of bias assessment: each study was assessed depending on the study type (RCT, non-RCT, cohort study, case series, case report).^20–23^ Two reviewers (S.Z., W.W.P.) independently applied respective tools, discrepancies were discussed with a third reviewer (J.W.S.) until consensus was achieved. The results of assessment depending on the proportion of “yes” answers were scored in the following way: ≤49% yes = high risk of bias, 50-69% yes = moderate risk of bias, ≥70% yes = low risk of bias.

### Statistical analysis

Patient and treatment characteristics are presented overall, and according to 60-day survival, using median (25th, 75th percentile) for continuous variables and frequency count (%) for nominal variables. Since the predominant variants of SARS-CoV-2, patient demographics, and disease severity changed over the study period, analyses to assess the association of patient and treatment characteristics with 60-day survival were performed using conditional logistic regression with 48 strata defined by calendar year (2020, 2021, 2022/2023), age (≤49, 50-59, 60-69, ≥70), sex (male, female), and WHO disease severity score (≤4, ≥5). For continuous variables, initial analyses were performed using a restricted cubic spline with 3 knots to test the linearity assumption. From these analyses, the linearity assumption was valid for all variables except the days since the last anti-CD20 dose. For the days since the last anti-CD20 dose, the log-odds of survival increased in a linear fashion up to 100 days and then remained relatively unchanged beyond 100 days. For this reason, the days since the last anti-CD20 dose was modeled using a linear term with the number of days truncated at 100 for those with values greater than 100. Separate conditional logistic regression models were fit for each patient and treatment characteristic with results summarized by presenting the odds ratio with corresponding 95% confidence interval. For these analyses, two-tailed p<0.05 was considered statistically significant. To further explore which characteristics had the greatest impact on 60-day survival, a machine learning model was developed with XGBoost chosen as the modeling framework because of its ability to handle missing data. For the XGBoost algorithm the outcome was 60-day survival and logarithmic loss was the evaluation metric to be minimized. Grid optimization of the learning algorithm was used to evaluate the effect of the learning rate (eta: 0.01, 0.1, or 0.3), maximum tree depth (3, 5, 7, or 10), minimum child weight (1, 3, or 5), subsample rate (80% or 100%), number of columns per tree (70% or 100%) and regularization (L1: 0 or 0.05; L2: 0.9 or 1). Using this search space, each combination of hyperparameter settings was trained using 5-fold cross-validation and using early stopping to determine the optimum number of trees. The final selected hyperparameters were eta=.1, maximum tree depth=3, minimum child weight=5, subsample rate=0.8, columns per tree=0.7, L1 regularization=0, L2 regularization=0.9 and the number of trees=35. Using the final hyperparameters, the model was retrained with training terminated after 35 iterations. A plot based on Shapley Additive Explanation (SHAP) values was constructed to quantify the relative importance of the variables in the final model. Receiver operating characteristic curve analysis was used to select a threshold based on the Youden index. Statistical analyses were performed using SAS statistical software, version 9.4 (SAS Institute Inc, Cary, NC) and R statistical software, version 4.3.2 (R Foundation for Statistical Computing, Vienna, Austria).

## Results

### Study selection and characteristics

The process of study selection for this meta-analysis is presented in the PRISMA flow diagram (Figure 1). One RCT^24^ with one eligible patient, one non-RCT^25^ with one patient, 11 cohort studies^10,11,26–34^ with a total of 268 eligible patients, 27 case series^35–60^ with a total of 253 eligible patients, and 45 case reports^61–105^ with a total of 47 patients met our inclusion criteria.

Thus, the final analysis cohort included 570 patients from 85 studies. Only subgroups of patients were included from cohort studies, RCT and non-RCT, thus limiting the scope of attainable analysis to individual-level and rendering the use of a control group unfeasible. This was due to the lack of large studies primarily focused on the outcomes of B-cell depleted patients transfused with CCP, as these patients were often included into a cohort of immunocompromised patients. Data for the studies included in this work were collected mostly during the high-volume phases of the pandemic and thus represent a challenging data collection environment for the investigators, so variables of interest were not always available. Counts of total number of individuals with available data per variable are presented in Table 1 and Table 2. Descriptive and exploratory analyses were performed on the synthesized IPD from all included studies.

**Table 1.** Patient characteristics.

| Characteristic | N* | Overall Summary** | 60-day survival** |  |
| --- | --- | --- | --- | --- |
|  |  |  | No | Yes |
| Overall | 570 |  | 77 (13.5%) | 493 (86.5%) |
| Calendar year of infection | 570 |  |  |  |
| 2020 |  | 246 (43.2%) | 37 (15.0%) | 209 (85.0%) |
| 2021 |  | 115 (20.2%) | 19 (16.5%) | 96 (83.5%) |
| 2022/2023 |  | 209 (36.7%) | 21 (10.0%) | 188 (90.0%) |
| Patient type | 567 |  |  |  |
| Autoimmune disorder |  | 114 (20.1%) | 3 (2.6%) | 111 (97.4%) |
| Hematologic malignancy/disorder |  | 450 (79.4%) | 73 (16.2%) | 377 (83.8%) |
| Solid organ transplantation |  | 3 (0.5%) | 0 (0.0%) | 3 (100.0%) |
| Age, years | 565 | 64 (53, 72) | 70 (63, 77) | 62 (52, 70) |
| Sex | 569 |  |  |  |
| Female |  | 232 (40.8%) | 14 (6.0%) | 218 (94.0%) |
| Male |  | 337 (59.2%) | 63 (18.7%) | 274 (81.3%) |
| Days since the last anti-CD20 dose | 393 | 58 (22, 123) | 26 (15, 79) | 60 (24, 127) |
| WHO disease severity score | 562 |  |  |  |
| ≤ 4 |  | 393 (69.9%) | 28 (7.1%) | 365 (92.9%) |
| ≥ 5 |  | 169 (30.1%) | 48 (28.4%) | 121 (71.6%) |
| Number of COVID-19 vaccine doses | 493 |  |  |  |
| 0 |  | 222 (45.0%) | 30 (13.5%) | 192 (86.5%) |
| 1 |  | 13 (2.6%) | 2 (15.4%) | 11 (84.6%) |
| 2 |  | 97 (19.7%) | 23 (23.7%) | 74 (76.3%) |
| 3 |  | 92 (18.7%) | 14 (15.2%) | 78 (84.8%) |
| 4 or more |  | 69 (14.0%) | 2 (2.9%) | 67 (97.1%) |
| Anti-spike Ab status | 341 |  |  |  |
| Negative |  | 306 (89.7%) | 48 (15.7%) | 258 (84.3%) |
| Positive |  | 35 (10.3%) | 6 (17.1%) | 29 (82.9%) |

\*Number of patients with information available. The number of patients with information available for non-survivors/survivors was N=77/488 for age; and N=46/347 for days since the last anti-CD20 dose.
\*\*Data are summarized using median (25<sup>th</sup>, 75<sup>th</sup>) for continuous variables and n (%) for categorical variables. The percentages presented in the overall summary correspond to column percentages, and the percentages presented in the summaries according to 60-day survival correspond to row percentages.
WHO, World Health Organization. Ab, antibody.

**Table 2.**
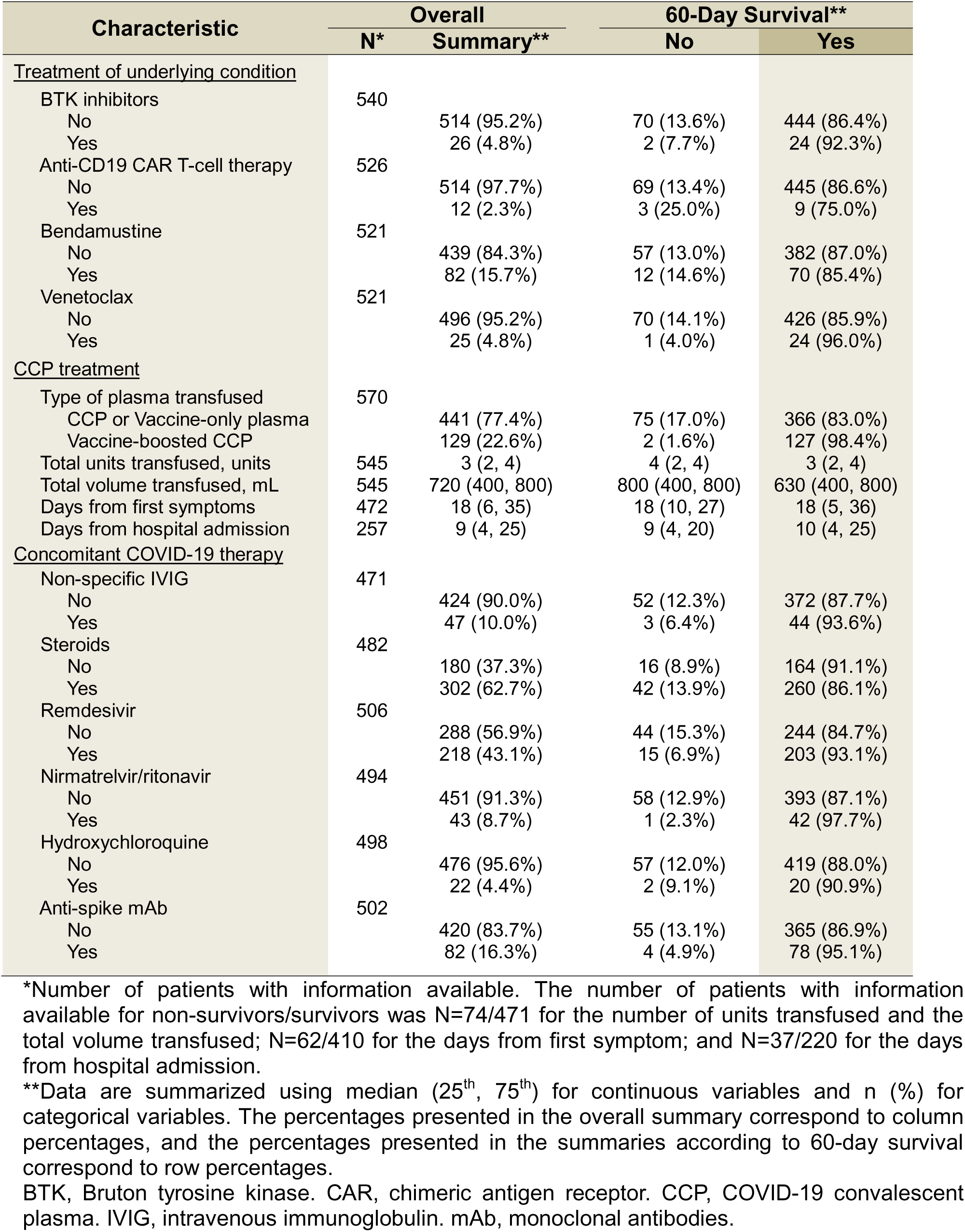
Treatment characteristics.

| Characteristic | N* | Overall<br>Summary** | 60-Day Survival** |  |
| --- | --- | --- | --- | --- |
|  |  |  | No | Yes |
| <u>Treatment of underlying condition</u> |  |  |  |  |
| BTK inhibitors | 540 |  |  |  |
| No |  | 514 (95.2%) | 70 (13.6%) | 444 (86.4%) |
| Yes |  | 26 (4.8%) | 2 (7.7%) | 24 (92.3%) |
| Anti-CD19 CAR T-cell therapy | 526 |  |  |  |
| No |  | 514 (97.7%) | 69 (13.4%) | 445 (86.6%) |
| Yes |  | 12 (2.3%) | 3 (25.0%) | 9 (75.0%) |
| Bendamustine | 521 |  |  |  |
| No |  | 439 (84.3%) | 57 (13.0%) | 382 (87.0%) |
| Yes |  | 82 (15.7%) | 12 (14.6%) | 70 (85.4%) |
| Venetoclax | 521 |  |  |  |
| No |  | 496 (95.2%) | 70 (14.1%) | 426 (85.9%) |
| Yes |  | 25 (4.8%) | 1 (4.0%) | 24 (96.0%) |
| <u>CCP treatment</u> |  |  |  |  |
| Type of plasma transfused | 570 |  |  |  |
| CCP or Vaccine-only plasma |  | 441 (77.4%) | 75 (17.0%) | 366 (83.0%) |
| Vaccine-boosted CCP |  | 129 (22.6%) | 2 (1.6%) | 127 (98.4%) |
| Total units transfused, units | 545 | 3 (2, 4) | 4 (2, 4) | 3 (2, 4) |
| Total volume transfused, mL | 545 | 720 (400, 800) | 800 (400, 800) | 630 (400, 800) |
| Days from first symptoms | 472 | 18 (6, 35) | 18 (10, 27) | 18 (5, 36) |
| Days from hospital admission | 257 | 9 (4, 25) | 9 (4, 20) | 10 (4, 25) |
| <u>Concomitant COVID-19 therapy</u> |  |  |  |  |
| Non-specific IVIG | 471 |  |  |  |
| No |  | 424 (90.0%) | 52 (12.3%) | 372 (87.7%) |
| Yes |  | 47 (10.0%) | 3 (6.4%) | 44 (93.6%) |
| Steroids | 482 |  |  |  |
| No |  | 180 (37.3%) | 16 (8.9%) | 164 (91.1%) |
| Yes |  | 302 (62.7%) | 42 (13.9%) | 260 (86.1%) |
| Remdesivir | 506 |  |  |  |
| No |  | 288 (56.9%) | 44 (15.3%) | 244 (84.7%) |
| Yes |  | 218 (43.1%) | 15 (6.9%) | 203 (93.1%) |
| Nirmatrelvir/ritonavir | 494 |  |  |  |
| No |  | 451 (91.3%) | 58 (12.9%) | 393 (87.1%) |
| Yes |  | 43 (8.7%) | 1 (2.3%) | 42 (97.7%) |
| Hydroxychloroquine | 498 |  |  |  |
| No |  | 476 (95.6%) | 57 (12.0%) | 419 (88.0%) |
| Yes |  | 22 (4.4%) | 2 (9.1%) | 20 (90.9%) |
| Anti-spike mAb | 502 |  |  |  |
| No |  | 420 (83.7%) | 55 (13.1%) | 365 (86.9%) |
| Yes |  | 82 (16.3%) | 4 (4.9%) | 78 (95.1%) |

\*Number of patients with information available. The number of patients with information available for non-survivors/survivors was N=74/471 for the number of units transfused and the total volume transfused; N=62/410 for the days from first symptom; and N=37/220 for the days from hospital admission.
\*\*Data are summarized using median (25<sup>th</sup>, 75<sup>th</sup>) for continuous variables and n (%) for categorical variables. The percentages presented in the overall summary correspond to column percentages, and the percentages presented in the summaries according to 60-day survival correspond to row percentages.
BTK, Bruton tyrosine kinase. CAR, chimeric antigen receptor. CCP, COVID-19 convalescent plasma. IVIG, intravenous immunoglobulin. mAb, monoclonal antibodies.

### Risk assessment

The results of the risk of bias assessment are presented in Tables S4-S8. Both RCT and non-RCT showed low risk of bias. Eight of 11 cohort studies showed low risk of bias, while only one cohort study showed high risk. Among 27 case series 21 were assessed to have low risk of bias, and 3 had high risk. When risk of bias assessment tool was applied to case reports, 40 of 45 studies showed low risk, and only one showed high risk of bias.

### Exploratory analyses of individual participant data

The final cohort included 570 patients who were B-cell depleted due to treatment of other diseases with anti-CD20 monoclonal antibodies and who were transfused with CCP for COVID-19. Overall, 60-day survival was 86.5% (493 of 570 patients), while 70.1% of patients achieved viral clearance (288 of 411 patients) and 64.3% had clinical improvement within 5 days of CCP transfusion (286 of 445 patients).

Patient and treatment characteristics of our synthesized cohort overall and according to 60-day survival outcome are presented in Tables 1 and 2 (overall characteristics with stratification based on calendar year are in Tables S1-S2). As expected, patients who survived 60-days were observed to be younger (median [Q1, Q3] age of 62 [52, 70] versus 70 [63, 77] years for those who did not survive 60-days). WHO disease severity scale utilized in the final cohort is presented in Table S9. Survival was better for patients with lower WHO disease severity (92.9% vs 71,6% for WHO ≤4 versus WHO ≥5), females (94.0% versus 81.3% for females versus males), and those treated later during the pandemic (85.0%, 83.5%, and 90.0% for calendar year 2020, 2021, and 2022/2023 respectively). Results from conditional logistic regression analyses assessing the association of the other patient and treatment characteristics with 60-day survival after controlling for calendar year, age, sex, and WHO disease severity are summarized in Table 3. After controlling for these variables, 60-day survival was found to be significantly higher for autoimmune disorders (OR=6.88, 95% CI 1.50 to 31.63, p=0.013), longer time since the last anti-CD20 dose (OR=1.16 per 10-day increase, 95% CI 1.04 to 1.29, p=0.007), and treatment with vaccine-boosted CCP (OR=9.49, 95% CI 2.01 to 44.82, p=0.005). Additionally, patient outcomes according to year of infection are presented in Table S3.

**Table 3.** Conditional logistic regression results.

| Characteristic | OR | 95% CI | p |
| --- | --- | --- | --- |
| Patient type |  |  | 0.013 |
| Autoimmune disorder | 6.88 | (1.50, 31.63) |  |
| Hematologic cancer/disorder | 1.00 | (reference) |  |
| Days since the last anti-CD20 dose† | 1.16 | (1.04, 1.29) | 0.007 |
| Number of COVID-19 vaccine doses† | 0.92 | (0.69, 1.22) | 0.566 |
| Anti-spike Ab status |  |  | 0.929 |
| Negative | 1.00 | (reference) |  |
| Positive | 0.95 | (0.31, 2.88) |  |
| <u>Treatment of underlying condition</u> |  |  |  |
| BTK inhibitors | 1.12 | (0.21, 6.11) | 0.984 |
| Anti-CD19 CAR T-cell therapy | 0.10 | (0.02, 0.66) | 0.016 |
| Bendamustine | 0.94 | (0.44, 2.05) | 0.884 |
| Venetoclax | 3.65 | (0.46, 29.23) | 0.223 |
| <u>CCP treatment</u> |  |  |  |
| Type of plasma transfused |  |  | 0.005 |
| CCP or Vaccine-only plasma | 1.00 | (reference) |  |
| Vaccine-boosted CCP | 9.49 | (2.01, 44.82) |  |
| Total units transfused, units | 0.93 | (0.78, 1.11) | 0.426 |
| Total volume transfused, mL | 0.98 | (0.81, 1.19) | 0.827 |
| Days from first symptoms | 1.01 | (1.00, 1.02) | 0.179 |
| Days from hospital admission | 1.00 | (0.99, 1.02) | 0.575 |
| <u>Concomitant COVID-19 therapy</u> |  |  |  |
| Non-specific IVIG | 1.82 | (0.48, 6.88) | 0.377 |
| Steroids | 0.95 | (0.45, 1.97) | 0.879 |
| Remdesivir | 1.31 | (0.66, 2.61) | 0.440 |
| Nirmatrelvir/ritonavir | 2.76 | (0.32, 23.74) | 0.354 |
| Hydroxychloroquine | 1.61 | (0.34, 7.76) | 0.550 |
| Anti-spike mAb | 2.17 | (0.66, 7.10) | 0.202 |

Each treatment characteristic was assessed separately using conditional logistic regression with strata defined by calendar year (2020, 2021, 2022/2023), age ( $\leq 49$ , 50-59, 60-69, $\geq 70$ ), sex (male, female), and WHO disease severity score ( $\leq 4$ , $\geq 5$ ). Results are summarized by presenting the odds ratio (OR) with 95% confidence interval (CI), where odds ratios greater than one correspond to higher odds of 60-day survival. For continuous variables, the odds ratio corresponds to a 10-day increase in the time since the last anti-CD20 dose, 1 dose increase in the number of COVID-19 vaccine doses, 1-unit increase for total units transfused, 225 ml increase for total volume transfused, and 1 day increase for days from first symptoms and days from hospital admission.
†For analysis purposes, patients with a value of $> 100$ for days since last anti-CD20 dose were assigned a value of 100; and patients with a value of $> 4$ for the number COVID-19 vaccine doses were assigned a value of 4.
Ab, antibody. BTK, Bruton tyrosine kinase. CAR, chimeric antigen receptor. CCP, COVID-19 convalescent plasma. IVIG, intravenous immunoglobulin. mAb, monoclonal antibodies.

To further explore which characteristics had the greatest impact on 60-day survival, a machine learning based model was developed using XGBoost. The model performance for the final model is shown in Figure S1. From the final model, the AUC was 0.916 (95% CI 0.887 to 0.946); and using a threshold of 0.83 or higher, the observed accuracy, sensitivity and specificity were 80.5% (459/570; 95% CI 77.0% to 83.7%), 78.7% (388/493; 95% CI 74.8% to 82.2%) and 92.2% (71/77; 95% CI 83.8% to 97.1%) respectively. A plot of the Shapley Additive Explanation (SHAP) values, demonstrating variable importance for the final XGBoost model, is shown in Figure 2. In order of importance, the top five variables were WHO disease severity ≥ 5, age, male sex, days since the last anti-CD20 dose, and treatment with vaccine-boosted CCP.

**Figure 2.**
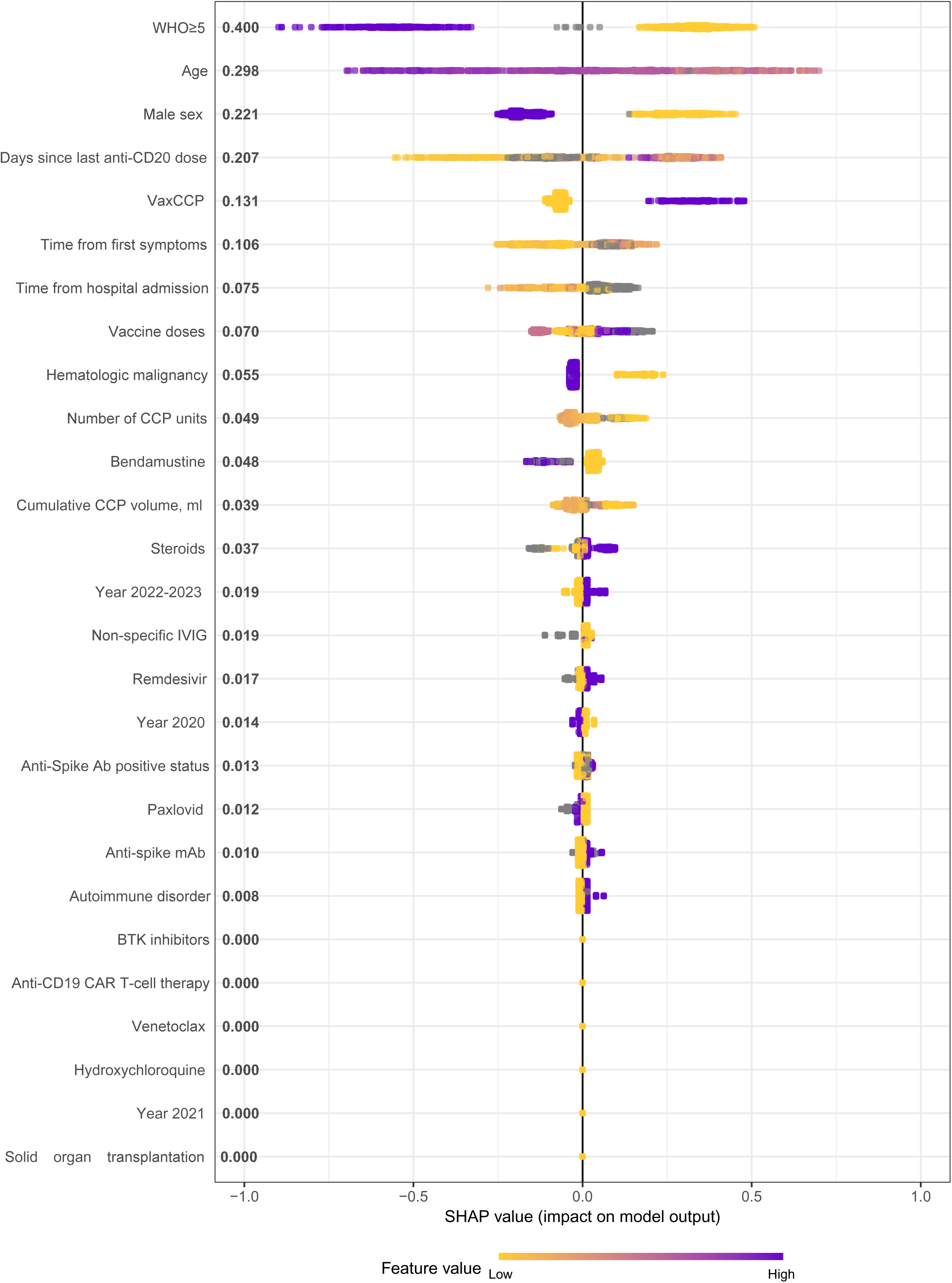
Shapley Additive Explanation (SHAP) values demonstrating variable importance, and direction, for the final XGBoost model. The SHAP value accounts for the interrelationship between the model features. Positive SHAP values indicate a higher likelihood of 60-day survival. Relative importance is presented after each variable with those having higher importance listed higher on the graph. Color is used to show the relative influence based on the value of the given variable, with yellow representing lower values and purple representing higher values. WHO, World Health Organization. VaxCCP, vaccine-boosted COVID-19 convalescent plasma. IVIG, intravenous immunoglobulin. mAb, monoclonal antibodies. BTK, Bruton tyrosine kinase. CAR, chimeric antigen receptor.

## Discussion

This study is the first systematic review and IPD meta-analysis to describe CCP efficacy in a clearly defined iatrogenic immunodeficiency, namely B-cell deficiency from anti-CD20 mAb therapy. This aspect of our study distinguishes it from the previous work in a more heterogenous population of immunocompromised patients. CCP transfusion in patients with B-cell deficiencies is akin to immunoglobulin replacement therapy, which is commonly used in antibody deficiencies.

This systematic review and IPD meta-analysis of B-cell depleted patients has four main findings. First, COVID-19 treatment with CCP in B-cell depleted patients was associated with a survival rate similar to the population of patients with different types of primary and secondary immunodeficiencies, transfused with CCP.^15^ Second, vaccine-boosted CCP, which likely had higher neutralizing antibody content, was associated with a survival benefit. Third, remdesivir did not show significant interaction with CCP when administered concomitantly in patients with different severities of COVID-19. Fourth, among B-cell depleted patients, underlying hematologic malignancies were associated with higher mortality when compared to patients who received B-cell depletion for autoimmune disorders.

In mid-2021, a novel vaccine-boosted convalescent plasma became available. Vaccine-boosted CCP contains 10 to 100 times more or higher antibody titers than the CCP used early in the pandemic.^106^ Vaccine-boosted CCP is derived from individuals who have both recovered from SARS-CoV-2 and received vaccination against SARS-CoV-2, or from vaccinated individuals after breakthrough infections. Previous research has shown higher titer CCP to be associated with survival benefit among patients not requiring mechanical ventilation before CCP transfusion^107^—a finding which was recapitulated in our results. Patients with WHO score ≤4 (did not require mechanical ventilation) had higher survival rates when transfused with vaccine-boosted CCP as opposed to conventional CCP.

Patients with autoimmune diseases had a higher survival rate (97.4%) than patients with hematologic malignancies (83.8%). After adjustment for year of infection, age, sex, and COVID-19 severity, autoimmune disorders were associated with better survival prognosis. This might be due to morbidity secondary to the underlying malignancies and the use of more aggressive concomitant immunosuppressive treatments in these patients. In a recent cohort study published after our systematic search was conducted, 92 patients with autoimmune disorders transfused with CCP had a similar survival rate of 97%.^108^ While another study reporting on outcomes of patients with hematologic diseases shows survival rates of 76.9% among the 81 B-cell depleted patients.^34^ That study included only hospitalized cases, however our combined group included both in- and out-patient individuals with COVID-19, thus potentially affecting discrepancies in survival rates.

Biological sex and age have been known risk factors affecting survival in not only immunocompromised, but also in the general population of COVID-19 patients^109^: our machine learning model shows that these associations remain the same in patients with B-cell depletion. Similarly, we confirm that B-cell depleted patient with milder presentations of COVID-19 (lower WHO severity score and lower requirements of ventilatory support) have better prognosis.

It is noteworthy that time from first symptom onset to first CCP transfusion had a wide range from 1 to 217 days, with a median of 18 days. These findings show that CCP remains effective in B-cell depleted patients even when administered late in COVID-19 course. Despite the protracted disease course, a majority of B-cell depleted patients achieved rapid clinical improvement (within 5 days of plasma transfusion) and were finally able to clear the virus. However, CCP characteristics such as number of units and total volume of transfused CCP did not significantly affect survival in our cohort. This observation might be attributed to protracted course of infection, non-standardized protocols of CCP transfusion with re-infusions at different time points of the disease and does not necessarily reflect lack of dose-response effect. Cases presented herein were challenging and did not always respond to standard treatment protocols, thus patient management was complex.

Our findings suggest that the time since last dose of anti-CD20 monoclonal antibodies represents a key factor associated with patient outcomes, as more recent anti-CD20 treatment was associated with higher mortality.

The COVID-19 pandemic has been so far associated with several different SARS-CoV-2 variants, including heterogenous wild-type genotypes early in the pandemic, supplanted by Alpha (B.1.1.7) and Beta (B.1.351) variants of concern (VOC) in early 2021, Delta VOC (B.1.617.2) in the middle of 2021, and Omicron VOC and its subvariants thereafter. In our study, patients with B-cell depletion who received CCP transfusion later in the pandemic (2022/2023) had better survival compared to patients receiving CCP before 2022. These differences in survival may be partly attributed to any of the following: advances in COVID-specific treatments and testing tools, emergence of biologically less virulent Omicron VOC, and also to the more precise identification of CCP units with high titer of neutralizing antibodies and earlier transfusion during later phases of the pandemic.

Studies have reported survival benefit in immunocompromised patients treated with remdesivir ^10 110^, while nirmatrelvir/ritonavir has shown to be associated with reduced mortality in non-immunocompromised population.^111^ However, our analysis did not show an interaction between remdesivir and CCP when administered concomitantly. Similarly, concomitant treatment with nirmatrelvir/ritonavir did not elicit significant interaction. We infer that this might be due different factors, such as protracted course of COVID-19 or CCP being already effective *per se* and masking the efficacy of remdesivir and nirmatrelvir/ritonavir.

Our study had some limitations. First, the exploratory analysis included data from lower tiers of evidence, such as case series and case reports (72 of 85 studies included). Although, these data cannot provide robust evidence of treatment effects, it can provide insights into the optimal use of an approved therapeutic with demonstrated safety and efficacy, such as CCP for treatment of B-cell depleted patients with COVID-19. Second, in our analysis we were limited to individual-level data analysis, since only two small cohort studies^10,26^ met our inclusion criteria on the group data level. Third, our synthesized cohort lacks control group that did not receive CCP. Last, we do not report antibody titers of transfused CCP, as some studies did not present titers due to lack of assay tools at the time, or tests were heterogenous among studies.

## Conclusion

This systematic review and IPD meta-analysis supports the efficacy profile of CCP as a treatment strategy in B-cell depleted patients with COVID-19 and identifies cases with better survival rates after treatment. As may be anticipated based on biological principles of antibody therapy^112^, these data show that the benefit of CCP was most apparent in patients who received CCP containing higher levels of antibodies – vaccine-boosted CCP.

## Data Availability

This secondary research did not generate original data, which remain available at the cited references.

## Author contributions

D.F., M.F., J.W.S. and M.J.J. conceived the study. D.F. created the data collection form. S.Z., D.F., D.C.H., A.J.M., J.W.S. abstracted individual patient data from the published literature. N.R., P.I., T.H., L.I.P., C.D., M.J., F.L., C.M-T., A.G., B.R., G.S., N.H., H.C. abstracted data from their own individual case series. S.Z., W.W.P., J.W.S. performed risk of bias assessment. D.R.S., R.E.C. performed statistical analyses. S.Z., D.F., D.R.S. wrote the first draft. All authors contributed to revising the manuscript and approved the final version of this manuscript.

