## Supplement for "Convalescent plasma treatment of B-cell depleted patients with COVID-19: systematic review and individual participant data meta-analysis"

This appendix has been provided by the authors to give readers additional information about their work.

### CONTENTS

**Table S1.** Patient characteristics according to year of infection. 3

**Table S2.** Treatment characteristics according to year of infection. 4

**Table S3.** Outcomes according to year of infection. 5

**Table S4.** Risk of bias assessment of randomized controlled trial. 6

**Table S5.** Risk of bias assessment of non-randomized controlled trial. 9

**Table S6.** Risk of bias assessment of cohort studies. 12

**Table S7.** Risk of bias assessment of case series. 13

**Table S8.** Risk of bias assessment of case reports. 15

**Table S9.** World Health Organization (WHO) disease severity scale for COVID-19. 17

**Table S10.** PRISMA Individual Participant Data meta-analysis checklist 2020. 18

**Figure S1.** Receiver operating characteristic curve from final XGBoost model for predicting 60-day survival. 23

**Table S1.** Patient characteristics according to year of infection.

| Characteristic | 2020 (N=246) | | 2021 (N=115) | | 2022/2023 (N=209) | |
|---|---|---|---|---|---|---|
| | N | Summary | N | Summary | N | Summary |
| Patient type | 245 | | 113 | | 209 | |
| Autoimmune disorder | | 30 (12.2%) | | 18 (15.9%) | | 66 (31.6%) |
| Hematologic malignancy/disorder | | 214 (87.3%) | | 95 (84.1%) | | 141 (67.5%) |
| Solid organ transplantation | | 1 (0.4%) | | 0 (0.0%) | | 2 (1.0%) |
| Age, years | 242 | 62 (52, 70) | 114 | 62 (53, 72) | 209 | 66 (56, 74) |
| Sex | 245 | | 115 | | 209 | |
| Female | | 98 (40.0%) | | 42 (36.5%) | | 92 (44.0%) |
| Male | | 147 (60.0%) | | 73 (63.5%) | | 117 (56.0%) |
| Days since the last anti-CD20 dose | 187 | 53 (23, 120) | 75 | 46 (13, 106) | 131 | 67 (25, 152) |
| Mechanical ventilation | 239 | 40 (16.7%) | 113 | 16 (14.2%) | 208 | 11 (5.3%) |
| WHO Disease Severity ≥ 5 | 241 | 93 (38.6%) | 113 | 39 (34.5%) | 208 | 37 (17.8%) |
| Number of COVID-19 vaccine doses | 197 | | 97 | | 199 | |
| 0 | | 155 (78.7%) | | 55 (56.7%) | | 12 (6.0%) |
| 1 | | 1 (0.5%) | | 9 (9.3%) | | 3 (1.5%) |
| 2 | | 26 (13.2%) | | 22 (22.7%) | | 49 (24.6%) |
| 3 | | 15 (7.6%) | | 9 (9.3%) | | 68 (34.2%) |
| 4 or more | | 0 (0.0%) | | 2 (2.1%) | | 67 (33.7%) |
| Anti-spike Ab status | 161 | | 77 | | 103 | |
| Negative | | 151 (93.8%) | | 67 (87.0%) | | 88 (85.4%) |
| Positive | | 10 (6.2%) | | 10 (13.0%) | | 15 (14.6%) |

Data are summarized using median ($25^{th}$, $75^{th}$) for continuous variables and n (%) for categorical variables.
WHO, World Health Organization. Ab, antibody.

**Table S2.** Treatment characteristics according to year of infection.

| Characteristic | 2020 (N=246) | | 2021 (N=115) | | 2022/2023 (N=209) | |
|---|---|---|---|---|---|---|
| | N | Summary | N | Summary | N | Summary |
| Treatment of underlying condition | | | | | | |
| BTK inhibitors | 239 | 5 (2.1%) | 103 | 4 (3.9%) | 198 | 17 (8.6%) |
| Anti-CD19 CAR T-cell therapy | 221 | 4 (1.8%) | 103 | 3 (2.9%) | 202 | 5 (2.5%) |
| Bendamustine | 220 | 37 (16.8%) | 100 | 14 (14.0%) | 201 | 31 (15.4%) |
| Venetoclax | 220 | 9 (4.1%) | 101 | 5 (5.0%) | 200 | 11 (5.5%) |
| CCP treatment | | | | | | |
| Type of plasma transfused | 246 | | 115 | | 209 | |
| CCP or Vaccine-only plasma | | 237 (96.3%) | | 105 (91.3%) | | 99 (47.4%) |
| Vaccine-boosted CCP | | 9 (3.7%) | | 10 (8.7%) | | 110 (52.6%) |
| Total units transfused, units | 234 | 3 (2, 4) | 105 | 3 (2, 4) | 206 | 3 (2, 4) |
| Total volume transfused, mL | 234 | 713 (400, 800) | 105 | 800 (450, 840) | 206 | 720 (360, 800) |
| Days from first symptoms* | 190 | 31 (17, 56) | 90 | 13 (7, 24) | 192 | 9 (2, 23) |
| Days from hospital admission* | 147 | 16 (6, 38) | 59 | 5 (2, 12) | 51 | 6 (4, 18) |
| Concomitant COVID-19 therapy | | | | | | |
| Non-specific IVIG | 182 | 25 (13.7%) | 90 | 9 (10.0%) | 199 | 13 (6.5%) |
| Steroids | 195 | 138 (70.8%) | 90 | 68 (75.6%) | 197 | 96 (48.7%) |
| Remdesivir | 201 | 81 (40.3%) | 104 | 51 (49.0%) | 201 | 86 (42.8%) |
| Nirmatrelvir/ritonavir | 199 | 0 (0.0%) | 97 | 0 (0.0%) | 198 | 43 (21.7%) |
| Hydroxychloroquine | 203 | 22 (10.8%) | 0 | 0 (0.0%) | 0 | 0 (0.0%) |
| Anti-spike mAb | 202 | 6 (3.0%) | 99 | 6 (6.1%) | 201 | 70 (34.8%) |

Data are summarized using median ($25^{th}$, $75^{th}$) for continuous variables and n (%) for categorical variables.
*Days to first plasma transfusion.
BTK, Bruton tyrosine kinase. CAR, chimeric antigen receptor. CCP, COVID-19 convalescent plasma. IVIG, intravenous immunoglobulin. mAb, monoclonal antibodies.

**Table S3.** Outcomes according to year of infection.

| Characteristic | 2020 (N=246) | | 2021 (N=115) | | 2022/2023 (N=209) | |
|---|---|---|---|---|---|---|
| | N | Summary | N | Summary | N | Summary |
| 60-day survival | 246 | 209 (85.0%) | 115 | 96 (83.5%) | 209 | 188 (90.0%) |
| Rapid improvement within 5 days | 177 | 107 (60.5%) | 79 | 47 (59.5%) | 189 | 132 (69.8%) |
| SARS-CoV-2 clearance | 194 | 132 (68.0%) | 100 | 75 (75.0%) | 117 | 81 (69.2%) |

Data are summarized using n (%).

**Table S4.** Risk of bias assessment of randomized controlled trial.

| Risk of bias for Denkinger et al. trial | Yes | No | Unclear | N/A |
|---|---|---|---|---|
| Bias related to selection and allocation | | | | |
| 1. Was true randomization used for assignment of participants to treatment groups? | ■ | □ | □ | □ |
| 2. Was allocation to treatment groups concealed? | ■ | □ | □ | □ |
| 3. Were treatment groups similar at the baseline? | □ | □ | ■ | □ |
| Bias related to administration of intervention/exposure | | | | |
| 4. Were participants blind to treatment assignment? | □ | ■ | □ | □ |
| 5. Were those delivering the treatment blind to treatment assignment? | □ | ■ | □ | □ |
| 6. Were treatment groups treated identically other than the intervention of interest? | ■ | □ | □ | □ |
| Bias related to assessment, detection and measurement of the outcome | | | | |
| 7. Were outcome assessors blind to treatment assignment? | | | | |
| Outcome 1 | □ | ■ | □ | □ |
| Outcome 2 | □ | ■ | □ | □ |
| Outcome 3 | □ | ■ | □ | □ |
| Outcome 4 | □ | ■ | □ | □ |
| Outcome 5 | □ | ■ | □ | □ |
| 8. Were outcomes measured in the same way for treatment groups? | | | | |
| Outcome 1 | ■ | □ | □ | □ |
| Outcome 2 | ■ | □ | □ | □ |

| | | | | |
|---|---|---|---|---|
| Outcome 3 | ■ | □ | □ | □ |
| Outcome 4 | ■ | □ | □ | □ |
| Outcome 5 | ■ | □ | □ | □ |
| 9. Were outcomes measured in a reliable way | | | | |
| Outcome 1 | ■ | □ | □ | □ |
| Outcome 2 | ■ | □ | □ | □ |
| Outcome 3 | ■ | □ | □ | □ |
| Outcome 4 | ■ | □ | □ | □ |
| Outcome 5 | ■ | □ | □ | □ |
| Bias related to participant retention | | | | |
| 10. Was follow up complete and if not, were differences between groups in terms of their follow up adequately described and analyzed? | | | | |
| Outcome 1 | ■ | □ | □ | □ |
| Outcome 2 | ■ | □ | □ | □ |
| Outcome 3 | ■ | □ | □ | □ |
| Outcome 4 | ■ | □ | □ | □ |
| Outcome 5 | ■ | □ | □ | □ |
| Statistical Conclusion Validity | | | | |
| 11. Were participants analyzed in the groups to which they were randomized? | | | | |
| Outcome 1 | ■ | □ | □ | □ |
| Outcome 2 | ■ | □ | □ | □ |

| | | | | |
|---|---|---|---|---|
| Outcome 3 | ■ | □ | □ | □ |
| Outcome 4 | ■ | □ | □ | □ |
| Outcome 5 | ■ | □ | □ | □ |
| 12. Was appropriate statistical analysis used? | | | | |
| Outcome 1 | ■ | □ | □ | □ |
| Outcome 2 | ■ | □ | □ | □ |
| Outcome 3 | ■ | □ | □ | □ |
| Outcome 4 | ■ | □ | □ | □ |
| Outcome 5 | ■ | □ | □ | □ |
| 13. Was the trial design appropriate and any deviations from the standard RCT design (individual randomization, parallel groups) accounted for in the conduct and analysis of the trial? | ■ | □ | □ | □ |

**Overall risk of bias: Low risk**

**Table S5.** Risk of bias assessment of non-randomized controlled trial.

| Risk of bias for Marconato et al. trial | Yes | No | Unclear | N/A |
|---|---|---|---|---|
| Bias related to temporal precedence | | | | |
| 1. Is it clear in the study what is the "cause" and what is the "effect" (i.e. there is no confusion about which variable comes first)? | ■ | □ | □ | □ |
| Bias related to selection and allocation | | | | |
| 2. Was there a control group? | □ | ■ | □ | □ |
| Bias related to confounding factors | | | | |
| 3. Were participants included in any comparisons similar? | □ | □ | □ | ■ |
| Bias related to administration of intervention/exposure | | | | |
| 4. Were the participants included in any comparisons receiving similar treatment/care, other than the exposure or intervention of interest? | □ | □ | □ | ■ |
| 5. Were there multiple measurements of the outcome, both pre and post the intervention/exposure? | | | | |
| Outcome 1 | ■ | □ | □ | □ |
| Outcome 2 | ■ | □ | □ | □ |
| Outcome 3 | ■ | □ | □ | □ |
| Outcome 4 | ■ | □ | □ | □ |
| Outcome 5 | ■ | □ | □ | □ |

| | | | | |
|---|---|---|---|---|
| 6. Were the outcomes of participants included in any comparisons measured in the same way? | | | | |
| Outcome 1 | ■ | ☐ | ☐ | ☐ |
| Outcome 2 | ■ | ☐ | ☐ | ☐ |
| Outcome 3 | ■ | ☐ | ☐ | ☐ |
| Outcome 4 | ■ | ☐ | ☐ | ☐ |
| Outcome 5 | ■ | ☐ | ☐ | ☐ |
| 7. Were outcomes measured in a reliable way? | | | | |
| Outcome 1 | ■ | ☐ | ☐ | ☐ |
| Outcome 2 | ■ | ☐ | ☐ | ☐ |
| Outcome 3 | ■ | ☐ | ☐ | ☐ |
| Outcome 4 | ■ | ☐ | ☐ | ☐ |
| Outcome 5 | ■ | ☐ | ☐ | ☐ |
| Bias related to participant retention | | | | |
| 8. Was follow-up complete and if not, were differences between groups in terms of their follow-up adequately described and analyzed? | | | | |
| Outcome 1 | ■ | ☐ | ☐ | ☐ |
| Outcome 2 | ■ | ☐ | ☐ | ☐ |
| Outcome 3 | ■ | ☐ | ☐ | ☐ |

| | | | | |
|---|---|---|---|---|
| Outcome 4 | ■ | ☐ | ☐ | ☐ |
| Outcome 5 | ■ | ☐ | ☐ | ☐ |
| Statistical Conclusion Validity | | | | |
| 9. Was appropriate statistical analysis used? | | | | |
| Outcome 1 | ☐ | ☐ | ☐ | ■ |
| Outcome 2 | ■ | ☐ | ☐ | ☐ |
| Outcome 3 | ■ | ☐ | ☐ | ☐ |
| Outcome 4 | ■ | ☐ | ☐ | ☐ |
| Outcome 5 | ■ | ☐ | ☐ | ☐ |
| **Overall risk of bias: Low risk** | | | | |

**Table S6.** Risk of bias assessment of cohort studies.

| Study | Risk of bias assessment tool questions | | | | | | | | | | | Overall risk |
|---|---|---|---|---|---|---|---|---|---|---|---|---|
| | Q1 | Q2 | Q3 | Q4 | Q5 | Q6 | Q7 | Q8 | Q9 | Q10 | Q11 | |
| Aiello et al, 2024 | N/A | N/A | Yes | Yes | No | Yes | Yes | Yes | No | Unclear | Yes | Moderate |
| Brown et al, 2022 | N/A | N/A | Yes | Yes | No | Yes | Unclear | Unclear | Unclear | N/A | Unclear | High |
| Destremau et al, 2024 | N/A | N/A | Yes | Yes | Yes | Yes | Yes | Yes | Yes | N/A | Yes | Low |
| Hueso et al, 2022 | N/A | N/A | Yes | Yes | Yes | Yes | Yes | Yes | Yes | N/A | Yes | Low |
| Ioannou et al, 2023 | N/A | N/A | Yes | Yes | Yes | Yes | Yes | Yes | Unclear | Unclear | Yes | Low |
| Lanza et al, 2022 | Yes | Yes | Yes | Yes | Yes | Yes | Yes | Yes | Yes | N/A | Yes | Low |
| Magyari et al, 2022 | N/A | N/A | Yes | Yes | No | Yes | Yes | Yes | Yes | N/A | Yes | Low |
| Richier et al, 2024 | N/A | N/A | Yes | Unclear | Unclear | Yes | Yes | Yes | Yes | N/A | Yes | Low |
| Thümmler et al, 2022 | No | Yes | Yes | Yes | Yes | Yes | Yes | Yes | Yes | N/A | Yes | Low |
| Weinbergerová et al, 2022 | Unclear | Yes | Yes | Yes | No | Yes | Yes | Unclear | Unclear | Unclear | Yes | Moderate |
| Weisser et al, 2022 | Yes | Yes | Yes | Unclear | Yes | Yes | Yes | Yes | Yes | Yes | Yes | Low |

Q1: Were the two groups similar and recruited from the same population? Q2: Were the exposures measured similarly to assign people to both exposed and unexposed groups? Q3: Was the exposure measured in a valid and reliable way? Q4: Were confounding factors identified? Q5: Were strategies to deal with confounding factors stated? Q6: Were the groups/participants free of the outcome at the start of the study (or at the moment of exposure)? Q7: Were the outcomes measured in a valid and reliable way? Q8: Was the follow up time reported and sufficient to be long enough for outcomes to occur? Q9: Was follow up complete, and if not, were the reasons to loss to follow up described and explored? Q10: Were strategies to address incomplete follow up utilized? Q11: Was appropriate statistical analysis used?
Overall risk of bias was scored in the following way: ≤49% yes = high risk of bias, 50-69% yes = moderate risk of bias, ≥70% yes = low risk of bias.

**Table S7.** Risk of bias assessment of case series.

| Study | Risk of bias assessment tool questions | | | | | | | | | | Overall risk |
|---|---|---|---|---|---|---|---|---|---|---|---|
| | Q1 | Q2 | Q3 | Q4 | Q5 | Q6 | Q7 | Q8 | Q9 | Q10 | |
| Bronstein et al, 2021 | Yes | Yes | Yes | Unclear | Unclear | Yes | Yes | Yes | Yes | N/A | Low |
| Cusi et al, 2021 | Yes | Yes | Yes | Unclear | Unclear | Yes | Yes | Yes | Yes | N/A | Low |
| D'Abramo et al, 2022 | Yes | Yes | Yes | Yes | Unclear | Yes | Yes | Yes | Yes | N/A | Low |
| Da Silva et al, 2023 | Yes | Unclear | Unclear | Unclear | Unclear | Yes | Yes | Yes | Unclear | N/A | High |
| Delgado-Fernández et al, 2022 | Yes | Yes | Yes | Yes | Yes | Yes | Yes | Yes | Yes | N/A | Low |
| Deveci et al, 2021 | Yes | Yes | Yes | Unclear | Unclear | Yes | Yes | Yes | No | N/A | Moderate |
| Erber et al, 2021 | Yes | Yes | Yes | Yes | Yes | Yes | Yes | Yes | Unclear | Yes | Low |
| Ferrari et al, 2021 | Yes | Yes | Yes | Yes | Unclear | Yes | Yes | Yes | Unclear | N/A | Low |
| Focosi et al, 2022 | Yes | Yes | Yes | Unclear | Unclear | No | Unclear | Yes | No | N/A | High |
| Franchini et al, 2022 | Yes | Yes | Yes | Unclear | Unclear | Yes | Yes | Yes | Yes | Yes | Low |
| Furlan et al, 2021 | Yes | Yes | Yes | Yes | Unclear | Yes | Yes | Yes | Unclear | N/A | Low |
| Gachoud 1 et al, 2022 | Yes | Yes | Yes | Yes | Unclear | Yes | Yes | Yes | No | Yes | Low |
| Gachoud 2 et al, 2022 | Yes | Yes | Yes | Yes | Unclear | No | No | Unclear | No | No | High |
| Gentile et al, 2024 | Yes | Yes | Yes | Yes | Yes | Yes | Yes | Yes | Yes | Yes | Low |
| Gharbharan et al, 2021 | Yes | Yes | Yes | Yes | Unclear | Yes | Yes | Yes | Unclear | N/A | Low |
| Hueso et al, 2020 | Yes | Yes | Yes | Yes | Yes | Yes | Yes | Yes | Unclear | N/A | Low |
| Huygens et al, 2023 | Yes | Yes | Yes | Yes | Yes | Yes | Yes | Yes | No | N/A | Low |
| Kenig et al, 2021 | Yes | Yes | Yes | Yes | Unclear | Yes | Yes | Yes | Yes | N/A | Low |
| Kluger et al, 2021 | Yes | Yes | Yes | Unclear | Unclear | Yes | Yes | Yes | No | N/A | Moderate |
| Kremer et al, 2021 | Yes | Yes | Yes | Unclear | Unclear | Yes | Yes | Yes | No | Yes | Low |
| Ljungquist et al, 2021 | Yes | Yes | Yes | Yes | Yes | Yes | Yes | Yes | Yes | Yes | Low |
| Martínez-Barranco et al, 2021 | Yes | Yes | Yes | Yes | Unclear | Yes | Yes | Yes | Yes | N/A | Low |
| Nissen-Meyer et al, 2023 | Yes | Unclear | Unclear | Yes | Yes | Yes | Yes | Yes | Yes | Yes | Low |
| Oliva et al, 2022 | Yes | Yes | Yes | Yes | Yes | Yes | Yes | Yes | Yes | N/A | Low |
| Prasad et al, 2021 | Yes | Yes | Yes | Unclear | Unclear | Yes | Yes | Yes | Unclear | N/A | Moderate |
| Ripoll et al, 2022 | Yes | Yes | Yes | Yes | Unclear | Yes | Yes | Yes | Yes | N/A | Low |
| Rüfenacht et al, 2022 | Yes | Yes | Yes | Yes | Yes | Yes | Yes | Yes | Unclear | N/A | Low |

Q1: Were there clear criteria for inclusion in the case series? Q2: Was the condition measured in a standard, reliable way for all participants included in the case series? Q3: Were valid methods used for identification of the condition for all participants included in the case series? Q4: Did the case series have consecutive inclusion of participants? Q5: Did the case series have complete inclusion of participants? Q6: Was there clear reporting of the demographics of the participants in the study? Q7: Was there clear reporting of clinical information of the participants? Q8: Were the outcomes or follow up results of cases clearly reported? Q9: Was there clear reporting of the presenting site(s)/clinic(s) demographic information? Q10: Was statistical analysis appropriate?
Overall risk of bias was scored in the following way: ≤49% yes = high risk of bias, 50-69% yes = moderate risk of bias, ≥70% yes = low risk of bias.

**Table S8.** Risk of bias assessment of case reports.

| Study | Risk of bias assessment tool questions | | | | | | | | Overall risk |
|---|---|---|---|---|---|---|---|---|---|
| | Q1 | Q2 | Q3 | Q4 | Q5 | Q6 | Q7 | Q8 | |
| Adedoyin et al, 2021 | Yes | Yes | Unclear | Unclear | Unclear | Yes | N/A | Yes | Moderate |
| Aviv et al, 2021 | Yes | No | Yes | Yes | Unclear | Yes | N/A | Unclear | Moderate |
| Baang et al, 2020 | Yes | Yes | Yes | Yes | No | Yes | N/A | Yes | Low |
| Baek et al, 2023 | Yes | Yes | Yes | Yes | Yes | Yes | Yes | Yes | Low |
| Balashov et al, 2021 | Yes | Yes | Yes | Yes | Yes | Yes | N/A | Yes | Low |
| Basheer et al, 2021 | No | No | Yes | Yes | Yes | Yes | N/A | Unclear | Moderate |
| Belcari et al, 2022 | Yes | Yes | Yes | Yes | Yes | Yes | N/A | Yes | Low |
| Bruiners et al, 2022 | Yes | Yes | Yes | Yes | Yes | Yes | N/A | Unclear | Low |
| Casarola et al, 2021 | Yes | Yes | Yes | Yes | Unclear | Yes | N/A | Yes | Low |
| Chen et al, 2024 | Yes | Yes | Yes | Yes | Yes | Yes | N/A | Yes | Low |
| Clark et al, 2020 | Yes | Yes | Yes | Yes | Yes | Yes | Yes | Yes | Low |
| Colombo et al, 2022 | Yes | Yes | Yes | Yes | Yes | Yes | N/A | Yes | Low |
| Garcia-Vidal et al, 2022 | Unclear | Yes | Yes | Yes | Unclear | No | N/A | Yes | Moderate |
| Gibson et al, 2021 | Yes | Yes | Yes | Yes | Yes | Yes | N/A | Yes | Low |
| Helleberg et al, 2020 | Unclear | Yes | Yes | Yes | Yes | Yes | N/A | Yes | Low |
| Honjo et al, 2020 | Yes | Yes | Yes | Yes | Yes | Yes | N/A | Yes | Low |
| Hughes et al, 2021 | Yes | Unclear | Yes | Yes | Yes | Yes | N/A | Yes | Low |
| Jassem et al. 2021 | Yes | Yes | Yes | Yes | Yes | Yes | N/A | Yes | Low |
| Karaolidou et al, 2021 | Yes | Yes | Yes | Yes | Yes | Yes | N/A | Yes | Low |
| Khatamzas et al, 2022 | Unclear | Yes | No | Unclear | Unclear | No | N/A | Unclear | High |
| Lancman et al, 2020 | Yes | Yes | Yes | Yes | Yes | Yes | N/A | Unclear | Low |
| Librizzi et al, 2024 | Yes | Yes | Yes | Yes | Unclear | Yes | N/A | Yes | Low |
| Malsy et al, 2020 | Yes | Yes | Yes | Yes | Yes | Yes | N/A | Unclear | Low |
| Martínez-Chinchilla et al, 2022 | Yes | Yes | Yes | Yes | Unclear | Yes | N/A | Yes | Low |
| Martinot et al, 2020 | Yes | Yes | Yes | Yes | Yes | Yes | N/A | No | Low |
| McKemey et al, 2021 | Yes | Yes | Yes | Yes | Yes | Yes | N/A | Yes | Low |
| Moore et al, 2020 | Yes | Unclear | Yes | Yes | Yes | Yes | N/A | Yes | Low |
| Moutinho-Pereira et al, 2021 | Yes | Yes | Yes | Yes | Yes | Yes | N/A | Yes | Low |
| Niu et al, 2020 | Yes | Yes | Yes | Yes | Yes | Yes | N/A | Yes | Low |

| | | | | | | | | | |
|---|---|---|---|---|---|---|---|---|---|
| Ordaya et al, 2022 | Yes | Yes | Yes | Yes | Unclear | Yes | N/A | Yes | Low |
| Ormazabal Vélez et al, 2021 | Yes | Yes | Yes | Yes | Yes | Yes | Yes | Yes | Low |
| Reuken et al, 2021 | Yes | Yes | Yes | Yes | Unclear | Yes | N/A | Yes | Low |
| Rnjak et al, 2021 | Yes | Yes | Yes | Yes | Yes | Yes | N/A | Yes | Low |
| Rodriguez-Pla et al, 2021 | Yes | Yes | Yes | Yes | Unclear | Yes | N/A | Yes | Low |
| Schenker et al, 2021 | Yes | Yes | Yes | Yes | Yes | Yes | N/A | Yes | Low |
| Sepulcri et al, 2021 | Yes | Yes | Yes | Yes | Unclear | Yes | N/A | Yes | Low |
| Seth-Smith et al, 2023 | Unclear | Yes | Yes | Yes | Yes | Yes | N/A | Yes | Low |
| Spinicci et al, 2022 | Yes | Yes | Yes | Yes | Yes | Yes | N/A | Yes | Low |
| Szwebel et al, 2021 | No | Yes | Yes | Yes | Yes | Yes | N/A | Yes | Low |
| Taha et al, 2021 | Yes | Yes | Yes | Yes | Unclear | Yes | N/A | Yes | Low |
| Tomisti et al, 2023 | Yes | Yes | Yes | Yes | Yes | Yes | Yes | Yes | Low |
| Villaseñor-Echavarri et al, 2023 | Yes | Yes | Yes | Yes | Yes | Yes | N/A | Yes | Low |
| Wright et al, 2021 | Yes | Yes | Yes | Yes | Yes | Yes | N/A | Yes | Low |
| Zhu et al, 2023 | Yes | Yes | Yes | Yes | Unclear | Yes | N/A | Yes | Low |
| Zimmerli et al, 2021 | Yes | Yes | Yes | Yes | Unclear | Yes | N/A | Yes | Low |

Q1: Were patient's demographic characteristics clearly described? Q2: Was the patient's history clearly described and presented as a timeline? Q3: Was the current clinical condition of the patient on presentation clearly described? Q4: Were diagnostic tests or assessment methods and the results clearly described? Q5: Was the intervention(s) or treatment procedure(s) clearly described? Q6: Was the post-intervention clinical condition clearly described? Q7: Were adverse events (harms) or unanticipated events identified and described? Q8: Does the case report provide takeaway lessons?

Overall risk of bias was scored in the following way: ≤49% yes = high risk of bias, 50-69% yes = moderate risk of bias, ≥70% yes = low risk of bias.

**Table S9.** World Health Organization (WHO) disease severity scale for COVID-19.

| WHO ordinal clinical severity scale | |
|---|---|
| 0 | No clinical or virological evidence of infection |
| 1 | Ambulatory, no activity limitation |
| 2 | Ambulatory, activity limitation |
| 3 | Hospitalized, no oxygen therapy |
| 4 | Hospitalized, oxygen mask or nasal prongs |
| 5 | Hospitalized, noninvasive mechanical ventilation or high-flow nasal cannula |
| 6 | Hospitalized, intubation and invasive mechanical ventilation (IMV) |
| 7 | Hospitalized, IMV + additional support such as pressors or extracardiac membranous oxygenation |
| 8 | Death |

**Table S10.** PRISMA Individual Participant Data meta-analysis checklist 2020.

| PRISMA-IPD Section/topic | Item No | Checklist item | Reported on page |
|---|---|---|---|
| **Title** | | | |
| Title | 1 | Identify the report as a systematic review and meta-analysis of individual participant data. | Title |
| **Abstract** | | | |
| Structured summary | 2 | Provide a structured summary including as applicable: | Abstract |
| | | **Background**: state research question and main objectives, with information on participants, interventions, comparators and outcomes. | |
| | | **Methods**: report eligibility criteria; data sources including dates of last bibliographic search or elicitation, noting that IPD were sought; methods of assessing risk of bias. | |
| | | **Results**: provide number and type of studies and participants identified and number (%) obtained; summary effect estimates for main outcomes (benefits and harms) with confidence intervals and measures of statistical heterogeneity. Describe the direction and size of summary effects in terms meaningful to those who would put findings into practice. | |
| | | **Discussion:** state main strengths and limitations of the evidence, general interpretation of the results and any important implications. | |
| | | **Other:** report primary funding source, registration number and registry name for the systematic review and IPD meta-analysis. | |
| **Introduction** | | | |
| Rationale | 3 | Describe the rationale for the review in the context of what is already known. | Introduction |
| Objectives | 4 | Provide an explicit statement of the questions being addressed with reference, as applicable, to participants, interventions, comparisons, outcomes and study design (PICOS). Include any hypotheses that relate to particular types of participant-level subgroups. | Introduction |
| **Methods** | | | |
| Protocol and registration | 5 | Indicate if a protocol exists and where it can be accessed. If available, provide registration information including registration number and registry name. Provide publication details, if applicable. | Protocol and registration |
| Eligibility criteria | 6 | Specify inclusion and exclusion criteria including those relating to participants, interventions, comparisons, outcomes, study design and characteristics (e.g. years when conducted, required minimum follow-up). Note whether these were applied at the study or individual level i.e. whether eligible participants were included (and | Eligibility criteria |

| | | | |
|---|---|---|---|
| | | ineligible participants excluded) from a study that included a wider population than specified by the review inclusion criteria. The rationale for criteria should be stated. | |
| Identifying studies - information sources | 7 | Describe all methods of identifying published and unpublished studies including, as applicable: which bibliographic databases were searched with dates of coverage; details of any hand searching including of conference proceedings; use of study registers and agency or company databases; contact with the original research team and experts in the field; open adverts and surveys. Give the date of last search or elicitation. | Information sources and search |
| Identifying studies - search | 8 | Present the full electronic search strategy for at least one database, including any limits used, such that it could be repeated. | Information sources and search |
| Study selection processes | 9 | State the process for determining which studies were eligible for inclusion. | Study selection and data collection processes |
| Data collection processes | 10 | Describe how IPD were requested, collected and managed, including any processes for querying and confirming data with investigators. If IPD were not sought from any eligible study, the reason for this should be stated (for each such study). | Study selection and data collection processes |
| | | If applicable, describe how any studies for which IPD were not available were dealt with. This should include whether, how and what aggregate data were sought or extracted from study reports and publications (such as extracting data independently in duplicate) and any processes for obtaining and confirming these data with investigators. | |
| Data items | 11 | Describe how the information and variables to be collected were chosen. List and define all study level and participant level data that were sought, including baseline and follow-up information. If applicable, describe methods of standardising or translating variables within the IPD datasets to ensure common scales or measurements across studies. | Study selection and data collection processes |
| IPD integrity | A1 | Describe what aspects of IPD were subject to data checking (such as sequence generation, data consistency and completeness, baseline imbalance) and how this was done. | Study selection and data collection processes |
| Risk of bias assessment in individual studies. | 12 | Describe methods used to assess risk of bias in the individual studies and whether this was applied separately for each outcome. If applicable, describe how findings of IPD checking were used to inform the assessment. Report if and how risk of bias assessment was used in any data synthesis. | Ethical Approval and Quality Assessment |

| | | | |
|---|---|---|---|
| Specification of outcomes and effect measures | 13 | State all treatment comparisons of interests. State all outcomes addressed and define them in detail. State whether they were pre-specified for the review and, if applicable, whether they were primary/main or secondary/additional outcomes. Give the principal measures of effect (such as risk ratio, hazard ratio, difference in means) used for each outcome. | Study selection and data collection processes |
| Synthesis methods | 14 | Describe the meta-analysis methods used to synthesise IPD. Specify any statistical methods and models used. Issues should include (but are not restricted to):<br>• Use of a one-stage or two-stage approach.<br>• How effect estimates were generated separately within each study and combined across studies (where applicable).<br>• Specification of one-stage models (where applicable) including how clustering of patients within studies was accounted for.<br>• Use of fixed or random effects models and any other model assumptions, such as proportional hazards.<br>• How (summary) survival curves were generated (where applicable).<br>• Methods for quantifying statistical heterogeneity (such as $I^2$ and $t^2$).<br>• How studies providing IPD and not providing IPD were analysed together (where applicable).<br>• How missing data within the IPD were dealt with (where applicable). | Study selection and data collection processes<br>&<br>Statistical analysis |
| Exploration of variation in effects | A2 | If applicable, describe any methods used to explore variation in effects by study or participant level characteristics (such as estimation of interactions between effect and covariates). State all participant-level characteristics that were analysed as potential effect modifiers, and whether these were pre-specified. | N/A |
| Risk of bias across studies | 15 | Specify any assessment of risk of bias relating to the accumulated body of evidence, including any pertaining to not obtaining IPD for particular studies, outcomes or other variables. | Ethical Approval and Quality Assessment |
| Additional analyses | 16 | Describe methods of any additional analyses, including sensitivity analyses. State which of these were pre-specified. | Statistical analysis |
| **Results** | | | |
| Study selection and IPD obtained | 17 | Give numbers of studies screened, assessed for eligibility, and included in the systematic review with reasons for exclusions at each stage. Indicate the number of studies and participants for which IPD were sought and for which IPD were obtained. For those studies where IPD were not available, give the numbers of studies and participants for which aggregate data were available. Report reasons for non-availability of IPD. Include a flow diagram. | Study selection and characteristics |

| | | | |
|---|---|---|---|
| Study characteristics | 18 | For each study, present information on key study and participant characteristics (such as description of interventions, numbers of participants, demographic data, unavailability of outcomes, funding source, and if applicable duration of follow-up). Provide (main) citations for each study. Where applicable, also report similar study characteristics for any studies not providing IPD. | Study selection and characteristics |
| IPD integrity | A3 | Report any important issues identified in checking IPD or state that there were none. | Study selection and characteristics |
| Risk of bias within studies | 19 | Present data on risk of bias assessments. If applicable, describe whether data checking led to the up-weighting or down-weighting of these assessments. Consider how any potential bias impacts on the robustness of meta-analysis conclusions. | Risk assessment & Tables S4-S8 |
| Results of individual studies | 20 | For each comparison and for each main outcome (benefit or harm), for each individual study report the number of eligible participants for which data were obtained and show simple summary data for each intervention group (including, where applicable, the number of events), effect estimates and confidence intervals. These may be tabulated or included on a forest plot. | Exploratory analysis of individual participant data |
| Results of syntheses | 21 | Present summary effects for each meta-analysis undertaken, including confidence intervals and measures of statistical heterogeneity. State whether the analysis was pre-specified, and report the numbers of studies and participants and, where applicable, the number of events on which it is based. | N/A |
| | | When exploring variation in effects due to patient or study characteristics, present summary interaction estimates for each characteristic examined, including confidence intervals and measures of statistical heterogeneity. State whether the analysis was pre-specified. State whether any interaction is consistent across trials. | |
| | | Provide a description of the direction and size of effect in terms meaningful to those who would put findings into practice. | |
| Risk of bias across studies | 22 | Present results of any assessment of risk of bias relating to the accumulated body of evidence, including any pertaining to the availability and representativeness of available studies, outcomes or other variables. | Risk assessment & Tables S4-S8 |
| Additional analyses | 23 | Give results of any additional analyses (e.g. sensitivity analyses). If applicable, this should also include any analyses that incorporate aggregate data for studies that do not have IPD. If applicable, summarise the main meta-analysis results following the inclusion or exclusion of studies for which IPD were not available. | Exploratory analysis of individual participant data |

| | | | |
|---|---|---|---|
| **Discussion** | | | |
| Summary of evidence | 24 | Summarize the main findings, including the strength of evidence for each main outcome. | Discussion |
| Strengths and limitations | 25 | Discuss any important strengths and limitations of the evidence including the benefits of access to IPD and any limitations arising from IPD that were not available. | Discussion |
| Conclusions | 26 | Provide a general interpretation of the findings in the context of other evidence. | Discussion |
| Implications | A4 | Consider relevance to key groups (such as policy makers, service providers and service users). Consider implications for future research. | Discussion |
| **Funding** | | | |
| Funding | 27 | Describe sources of funding and other support (such as supply of IPD), and the role in the systematic review of those providing such support. | Funding |

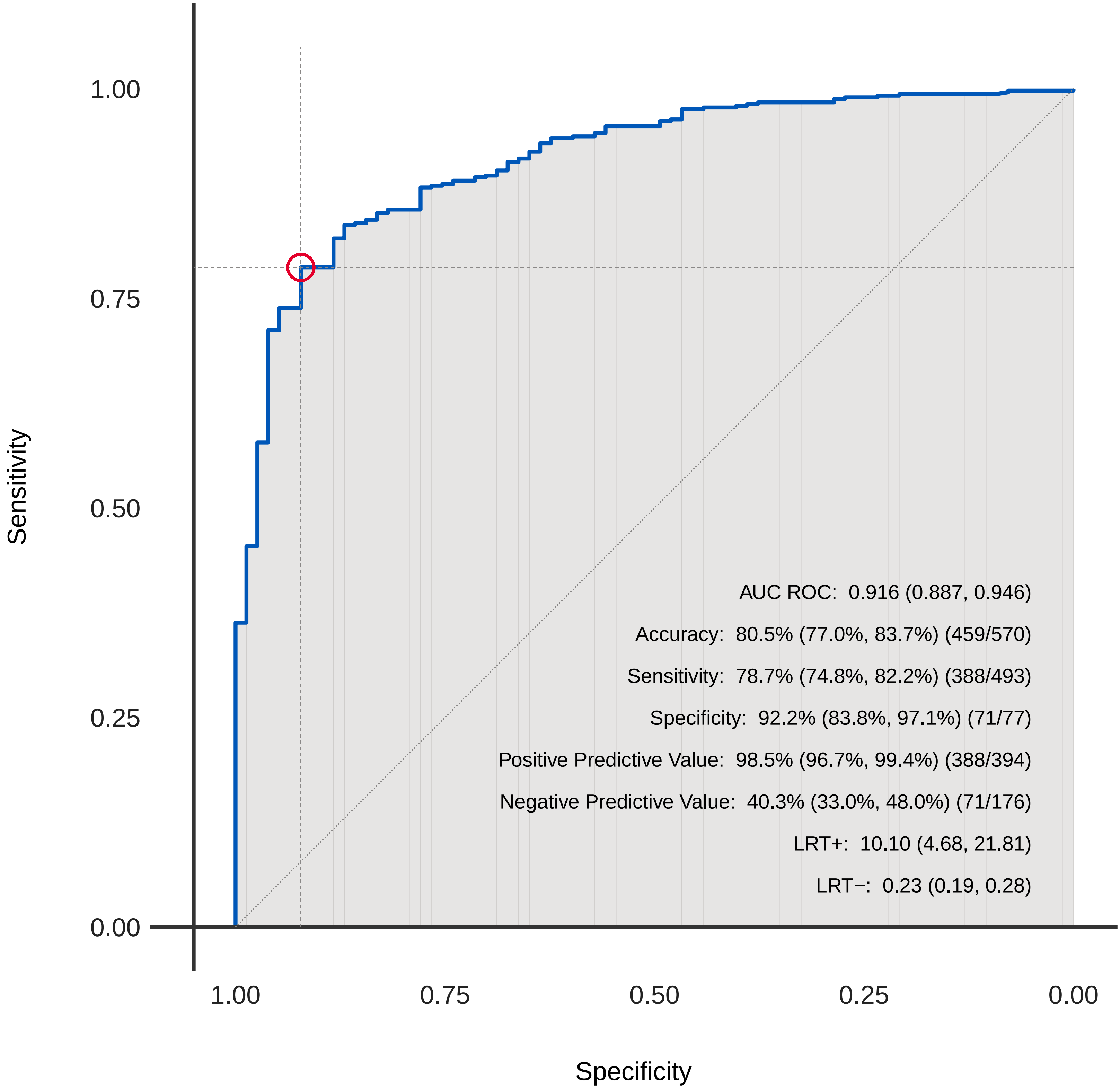

**Figure S1.** Receiver operating characteristic curve from final XGBoost model for predicting 60-day survival. Performance is summarized at a threshold of 0.83 or higher, which was chosen based on the Youden index. AUC, area under the receiver operating characteristic curve; LRT, likelihood ratio test.
